# Population structure and antimicrobial resistance patterns of *Salmonella* Typhi and Paratyphi A amid a phased municipal vaccination campaign in Navi Mumbai, India

**DOI:** 10.1101/2023.02.23.23286256

**Authors:** Kesia Esther Da Silva, Kashmira Date, Nilma Hirani, Christopher LeBoa, Niniya Jayaprasad, Priyanka Borhade, Joshua Warren, Rahul Shimpi, Seth Ari Sim-Son Hoffman, Matthew Mikoleit, Pankaj Bhatnagar, Yanjia Cao, Shanta Dutta, Stephen P Luby, Jason R. Andrews

## Abstract

We performed whole genome sequencing of 174 *Salmonella* Typhi and 54 *Salmonella* Paratyphi A isolates collected through prospective and retrospective surveillance in the context of a phased typhoid conjugate vaccine introduction in Navi Mumbai, India. We investigate the temporal and geographical patters of emergence and spread of antimicrobial resistance. Additionally, we evaluated the relationship between the spatial distance between households and genetic clustering of isolates using hierarchical Bayesian models. Most isolates were non-susceptible to fluoroquinolones, with nearly 20% containing ≥3 mutations in the quinolone resistance determining region, conferring high-level resistance. Two H58 isolates carried an IncX3 resistance plasmid containing *bla*_SHV-12_, associated with ceftriaxone resistance, suggesting that the ceftriaxone-resistant *S*. Typhi isolates from India have evolved independently on multiple occasions. Among *S*. Typhi isolates, we identified two main clades circulating in Navi Mumbai (2.2 and 4.3.1 [H58]); 2.2 isolates were closely related following a single introduction around 2007, whereas H58 isolates had been introduced multiple times to the city. Increasing geographic distance between isolates was strongly associated with genetic clustering (OR 0.72 per km; 95% CrI: 0.66-0.79). This effect was seen for distances up to 5 km (OR 0.65 per km; 95% CrI: 0.59-0.73) but was not seen for distances beyond 5 km (OR 1.02 per km; 95% CrI: 0.83-1.26). Our findings indicate that *S*. Typhi was repeatedly introduced into Navi Mumbai and then spread locally, with strong evidence of spatial-genetic clustering. In addition to vaccination, local interventions to improve water and sanitation will be critical to interrupt transmission.

## Introduction

Enteric fever, caused by *Salmonella enterica* serovars Typhi (*S*. Typhi) and Paratyphi A (*S*. Paratyphi A), is an acute febrile illness that remains one of the most critical infectious diseases globally (1). An estimated 11 million cases of typhoid fever and 3.5 million cases of paratyphoid fever occur worldwide each year, causing over 100,000 deaths, predominantly in low- and middle-income countries (2). Global data suggest the majority of the reported enteric fever morbidity and mortality take place in endemic regions of South Asia, Southeast Asia, and Africa (3). India is believed to have the largest number of typhoid cases in the world, and increasing prevalence of antimicrobial-resistant typhoid in the country is a major public health threat (4).

A particular multi-drug resistant (MDR) *S*. Typhi clone resistant to resistance to ampicillin, chloramphenicol, and trimethoprim-sulfamethoxazole, known as haplotype H58 or 4.3.1, has emerged over recent decades and now is prevalent across South and Southeast Asia and parts of Africa (5,6). Phylogenetic analysis indicated that South Asia might be the site of the original emergence of the 4.3.1 genotype (6). The emergence and spread of MDR *S*. Typhi led to increased reliance on fluoroquinolones for typhoid treatment (7); however, over the past 15 years, fluoroquinolone non-susceptible (FQNS) *S*. Typhi have become dominant throughout South Asia (8). More recently, the emergence of H58 *S*. Typhi ‘triple mutants’ harboring three quinolone resistance-determining regions (QRDR) mutations, associated with high-level fluoroquinolone resistance, now appear to be dominant in India (9). In the view of the high prevalence of FQNS *S*. Typhi, third generation cephalosporins are increasingly used for treatment of typhoid (10). However, reports of emergence of third generation cephalosporin-resistant *S*. Typhi have been described in numerous countries including India, posing a threat in future use of this drug for typhoid treatment (11–13).

The World Health Organization (WHO) recommends that typhoid conjugate vaccines (TCV) be used in settings with high typhoid burden or high prevalence of antimicrobial-resistant *S*. Typhi (14). India has not yet introduced TCVs nationally; however, in 2018, Navi Mumbai, a metropolitan city near Mumbai, introduced TCVs in half of its administrative areas, with a plan for providing vaccines to the other half that has been delayed due to the COVID-19 pandemic (15). To better understand the structure of the circulating pathogen population within an endemic area and after TCV introduction, we sequenced the genomes of 174 *S*. Typhi and 54 *S*. Paratyphi A isolates collected from Navi Mumbai between 2018 and 2021.

## Methods

### Setting and study population

Navi Mumbai is a city with a population of 1.12 million, including an estimated 129,500 children age 0-6 years (16). In September of 2018, a vaccine campaign was conducted in a randomly selected 11 of 22 urban health post (UHP) areas (designated phase 1), to children ages 9 months to 14 years. Coverage of the campaign was estimated to be 71% among age-eligible children living in these communities (15). The original plan was for the remaining communities to receive vaccination beginning 2 years later (phase 2); however, due to the COVID-19 pandemic, the second vaccine campaign has been delayed. To evaluate the effectiveness of the vaccine campaign, we performed prospective surveillance at six hospitals between June, 2018 and March, 2021. We recruited children with suspected enteric fever and performed blood culture for consenting participants (15). Additionally, culture-confirmed typhoid cases from a large private laboratory in the city were recruited into the study. All culture-confirmed cases from the parent study were eligible for inclusion in this genomic epidemiology study.

### Bacterial identification and antimicrobial susceptibility testing

*S*. Typhi and *S*. Paratyphi A were identified by biochemical profile and serotyping, and later confirmed by whole genome sequencing. Antimicrobial susceptibility to ampicillin, co-trimoxazole, chloramphenicol, ciprofloxacin and ceftriaxone was determined using the disk diffusion method (Oxoid, Thermo Scientific, MA, USA). All zone diameters were interpreted according to EUCAST v8.0 clinical breakpoints.

### Whole-genome sequencing

Genomic DNA was extracted using Promega Wizard® Genomic DNA Purification Kits (Promega Corporation, UK). Whole-genome sequencing (WGS) was performed at Genotypic Technology Pvt Ltd (Bangalore, India) using the Illumina Hiseq X Ten platform (Illumina, San Diego, CA, USA) to generate paired-end reads of 100–150 bp in length. Sequence data quality was checked using FastQC v0.11.9 to remove low quality reads (17). We summarized all quality indicators using MultiQC v1.7 (18). Species identification was confirmed with Kraken2 (19), and the *Salmonella in silico* Typing Resource (SISTR) was used for WGS-based serotyping (20). Raw data assembly was achieved using SPADES assembler. Short Read Sequence Typing for Bacterial Pathogens (SRST2) (21) was used to map known alleles and identify MLSTs directly from reads according to the *Salmonella enterica* MLST scheme (https://pubmlst.org/salmonella/).

### Mapping and SNP analysis

Paired-end Illumina reads were mapped to the *S*. Typhi CT18 (accession no. AL513382) reference chromosome sequence using RedDog mapping pipeline v1beta.11 (https://github.com/katholt/reddog). RedDog uses Bowtie2 v2.4.1 (22) to map reads to the reference genome, and SAMtools v1.10 (23) to identify SNPs that have a phred quality score above 30, and to filter out those SNPs supported by less than five reads, or with 2.5x the average read depth that represent repeated sequences, or those that have ambiguous base calls. For each SNP that passes these criteria in any one isolate, consensus base calls for the SNP locus were extracted from all genomes mapped, with those having phred quality scores under 20 being treated as unknown alleles and represented with a gap character.

Chromosomal SNPs with confident homozygous calls (phred score above 20) in >95% of the genomes mapped (representing a ‘soft’ core genome) were concatenated to form an alignment of alleles using the RedDog python script parseSNPtable.py with parameters -m cons, aln and –c 0.95 and SNPs called in prophage regions and repetitive sequences (354 kb; ∼7.4% of bases) in the CT18 reference chromosome, as defined previously (24) were excluded. SNPs occurring in recombinant regions were detected by Gubbins v2.4.1 (25) and excluded. The SNP data were used to assign all isolates to previously defined genotypes according to an extended *S*. Typhi genotyping framework using the GenoTyphi python script (https://github.com/katholt/genotyphi) (24).

To characterize and analyze the genomes of the 54 *S*. Paratyphi A strains, a similar bioinformatic process was adopted using *S*. Paratyphi A AKU_12601[27] (accession no: FM200053) as the reference genome to create an alignment with another selected 108 Indian isolates from previous studies (26,27). Genotypes were assigned according to an *S*. Paratyphi A genotyping framework (28) using the Paratype script (https://github.com/CHRF-Genomics/Paratype).

### Analysis of spatial distance and genetic clustering

We sought to evaluate whether spatial distance was associated with genetic clustering, hypothesizing that pairs of isolates from individuals whose homes are closer to one another are more likely to be genetically similar compared with pairs of individuals who live further from one another. One of the obstacles to testing this hypothesis is that individuals are represented across multiple pairs, such that the spatial and genetic relatedness of each pairwise comparison is not independent. To overcome this, we leveraged a recently developed hierarchical Bayesian modeling approach that accommodates this type of network dependence, along with spatial correlation, by incorporating spatially correlated individual-level random effects parameters into the regression framework (29). This approach has been implemented in a R package (*GenePair*). We evaluated an outcome of genetic clustering, defined as pairs of isolates that have fewer than 6 single nucleotide polymorphisms between them, following earlier literature (30), and in sensitivity analyses we examined other thresholds (3 SNPs, 12 SNPs). We extended this model using splines to investigate whether the relationship between spatial distance and clustering varied as a function of distance. We fit conventional generalized additive models, using the *mgcv* package in R, to first visualize the relationship between spatial distance and clustering in models not accounting for network dependence and spatial correlation, and then refit the *GenePair* models introducing splines at different points. Finally, to test whether isolates from individuals in vaccination clusters were more or less likely to genetic cluster compared with isolates from individuals in non-vaccination clusters, we fit models introducing dummy variables for pairs in which none, one, or both individuals were in a vaccination cluster. Each model was run where we collected 100,000 iterations from the algorithm; the first 50,000 were discarded prior to convergence of the model and the remainder were thinned by a factor of 10 prior to summarizing the posterior distribution for each parameter in order to reduce posterior autocorrelation. We report the posterior median and quantile-based 95% credible intervals (CrI) for each estimate. Models were checked visually and by using Geweke’s Z score to ensure convergence.

### Phylogenetic analyses

Maximum likelihood (ML) phylogenetic trees were inferred from the chromosomal SNP alignments using RAxML v8.2.10 (31) (command raxmlHPC-PTHREADS). A generalized time-reversible model and a Gamma distribution was used to model site-specific rate variation (GTR+ Γ substitution model; GTRGAMMA in RAxML) with 100 bootstrap pseudo replicates used to assess branch support for the ML phylogeny. We selected the single tree with the highest likelihood score as the best tree. The resulting phylogenies were visualized and annotated using the iTOL v5 online version (32).

### Temporal and Phylogeography analysis

To investigate dates of emergence and geographical transfers, we inferred timed phylogenies using temporally representative samples. To estimate evolutionary rates and times of common ancestry of isolates we used *treedater* R package with an uncorrelated relaxed molecular clock and repeating the procedure 100 times (33). Finally, we reconstructed the ancestral state of nodes using the maximum parsimony approach with *Phangorn* R package (https://www.rdocumentation.org/packages/phangorn/versions/2.8.1), considering events with a location probability of >0.5 between connected nodes. We considered a geographic transfer when the most probable location between two connected nodes (or between a node and a tip) differed, and we considered the time window of transfer as the date range between the nodes (or between the node and tip). The geospatial transmissions of lineage strains from the phylogeographic reconstructions were analyzed and visualized using ArcMap 10.7.1 (https://desktop.arcgis.com/en/arcmap/).

### Resistome analysis

ARIBA (Antimicrobial Resistance Identifier by Assembly) v2.10.0 and CARD database v1.1.8 (https://card.mcmaster.ca/home) were used to investigate antimicrobial resistance gene content. Point mutations in the quinolone resistance determining region (QRDR) of the DNA-gyrase *gyrA/B* and topoisomerase-IV *parC/E* genes, associated with reduced susceptibility to fluoroquinolones and quinolone resistance genes (*qnrS*) were also detected using ARIBA. Isolates were defined as being MDR if resistance genes were detected by ARIBA in the β-lactams, Trimethoprim, Sulphonamides, and Chloramphenicol classes. Plasmid replicons were identified using ARIBA and the PlasmidFinder database (30).

### Data availability

Details and accession numbers of sequence data included in our analysis are provided in the supplement (Tables S1-S2).

### Ethics statement

The study was approved by the MGM New Bombay Hospital IRB, Vashi, India; Institutional Ethics Committee, Indian Council of Medical Research – National Institute of Cholera and Enteric Diseases (No. A-1/2020-IEC); WHO Research Ethics Review Committee (ERC.0002923); and Stanford University IRB (IRB-39627).

## Results

### The population structure of S. Typhi isolates *in Navi Mumbai*

A total of 174 *S*. Typhi isolates were available for genome sequencing. Among these, 33 (19%) were collected before the vaccine campaign began and the remainder (141) were collected after. Genotype analysis showed that the pathogen population structure in Navi Mumbai is diverse, with 10 distinct genotypes identified (Figure 1a). Genotype 4.3.1 (H58) was dominant, accounting for over half (97/174; 55.7%) of all isolates. The major sublineages of H58 (lineage I and lineage II) were present among our isolates, with lineage II (genotype 4.3.1.2) comprising the majority (75/97; 77%) of H58 isolates. The second most prevalence Genotype was 2.2 (33%; 58/174), and isolates from this clade were closely related. The two main clades circulating in Navi Mumbai (genotypes 2.2 and 4.3.1.2) were see throughout Navi Mumbai, and we identified very little spatial aggregation of genotypes (Figure 1).

**Figure 1.**
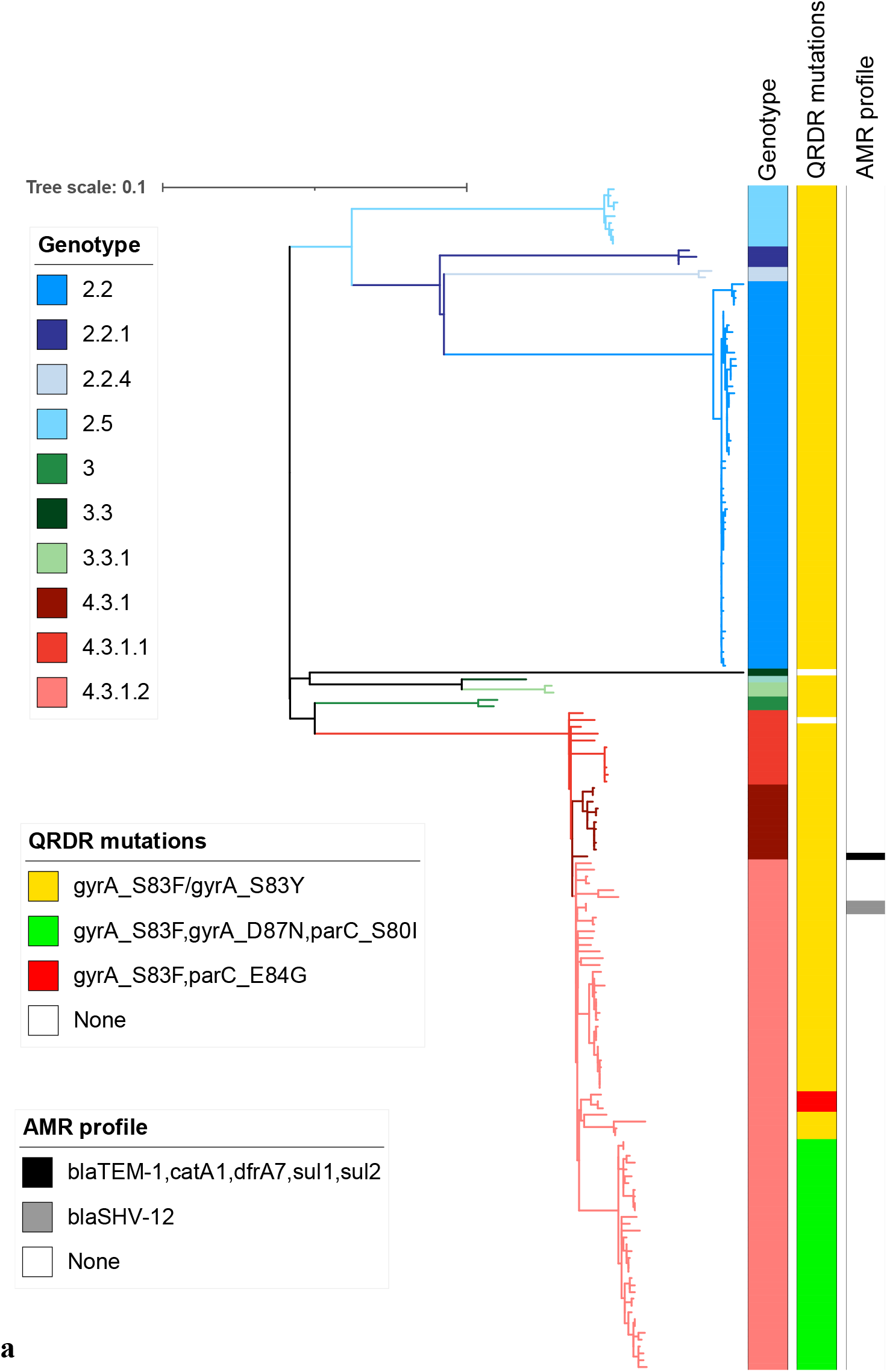
Navi Mumbai *Salmonella* Typhi population structure. **(a)** Maximum likelihood tree of 174 *S*. Typhi isolates from Navi Mumbai. Branch colors indicate the lineages. The scale bar indicates nucleotide substitutions per site.

**Figure 1(b).**
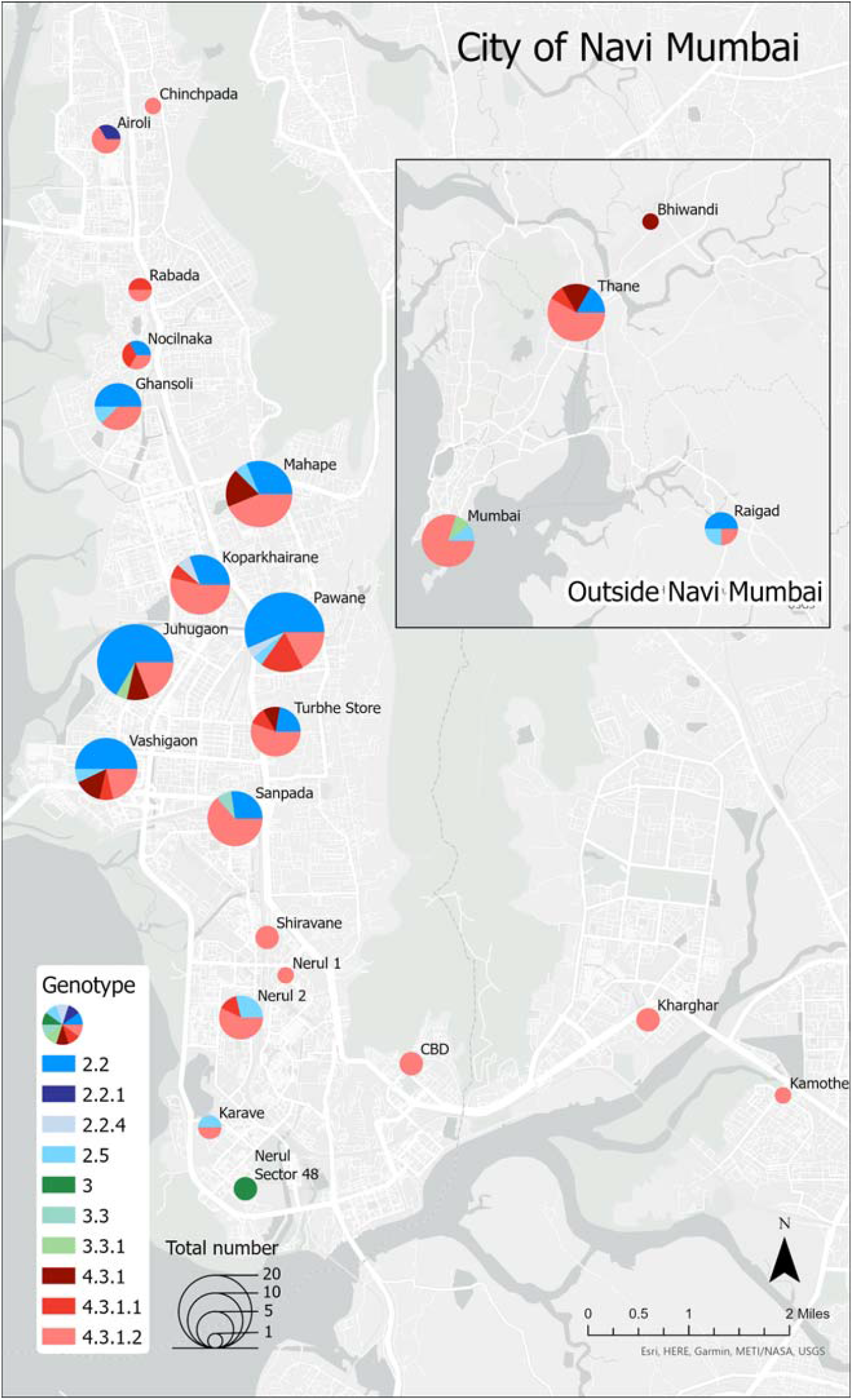
Map showing the distribution of genotypes by region.

### Antimicrobial Resistance characterization

Most *S*. Typhi isolates (98.3%) and all *S*. Paratyphi A were susceptible to traditional first-line antibiotics co-trimoxazole, ampicillin and chloramphenicol. We identified antimicrobial resistance genes to any of these three antibiotics in three isolates and a MDR profile (*bla*_TEM-1_, *catA1, dfrA7, sul1, sul2*) was observed in only one *S*. Typhi isolate, which belonged to 4.3.1 genotype. Two H58 (4.3.1.2) isolates were carrying an IncX3 resistance plasmid containing *bla*_SHV-12_, associated with ceftriaxone resistance. The majority of *S*. Typhi (99.4%) isolates and all the *S*. Paratyphi A isolates were fluoroquinolone non-susceptible (FQNS), primarily due to mutations in *gryA, gyrB, parC*, and *parE* (Figure 1a; Figure 3). Among our *S*. Typhi isolates, 34 (19.5%) were ‘triple mutants’ (Figure 1a), which are associated with high-level resistance to fluoroquinolones (34). All of the triple mutants were in H58 lineage II (4.3.1.2). Azithromycin resistance, conferred by *acrB* mutations (R717Q and R717L), was identified in only one *S*. Paratyphi A isolate, from genotype 2.4.3.

### Spatial distance and genetic clustering

In hierarchical Bayesian models, we found that increasing geographic distance between isolates was strongly associated with genetic clustering (OR 0.72 per km; 95% CrI: 0.66-0.79), using the 6 SNP threshold (**Table 1**). This effect was seen for distances up to 5 km (OR 0.65 per km; 95% CrI: 0.59-0.73) but was not seen for distances beyond 5 km (OR 1.02 per km; 95% CrI: 0.83-1.26). We observed no significant differences between the coefficients using shorter thresholds (e.g., 0-1 km vs 0-5 km). The relationship between spatial distance and genetic clustering was robust to the SNP distance threshold: OR per km of 0.69 (95% CrI: 0.62-0.76) for SNP threshold of 3 and OR per km of 0.79 (95% CrI: 0.74-0.84) for SNP threshold of 12. There was a non-significant reduction in odds of clustering for pairs of isolates in vaccination communities compared with pairs in non-vaccination communities (OR 0.42; 95% CrI: 0.11-1.43) or mixed pairs compared with pairs in non-vaccination communities (OR 0.73; 95% CrI: 0.19-2.54).

**Table 1.** Summary of results from spatial distance and genetic clustering analysis.

|  | aOR | 95% CrI |
| --- | --- | --- |
| **6 SNP threshold** |  |  |
| Distance | 0.72 | (0.66-0.79) |
| **6 SNP threshold with spline** |  |  |
| Distance (per km, up to 5 km) | 0.65 | (0.59-0.73) |
| Distance (per km, >5 km) | 1.02 | (0.83-1.26) |
| **3 SNP threshold** |  |  |
|  | 0.69 | (0.62-0.76) |
| **12 SNP threshold** |  |  |
|  | 0.79 | (0.74-0.84) |
| **6 SNP threshold with vaccination community** |  |  |
| Both in vaccine area vs both in non-vaccine area | 0.41 | (0.02-5.90) |
| One in each area versus both in non-vaccine area | 0.79 | (0.15-3.09) |

aOR – adjusted odds ratio.
CrI – credible interval.

### Intra-country transmission within India

To provide context for the genomes from Navi Mumbai and better understand temporal and spatial distribution of lineages, we constructed a whole genome phylogeny including 1,357 additional *S*. Typhi genomes previously sequenced from 17 cities in India (S1 Figure). Within 4.3.1 lineage, there was no tight clustering observed among the Navi Mumbai isolates. Instead, our study isolates manly clustered with previously sequenced isolates from neighboring Mumbai, indicating frequent transmission of 4.3.1 isolates between these cities. In comparison, all 2.2 isolates from Navi Mumbai were very closely related, and no clustering was observed with those from other cities.

### Evolutionary history of S. Typhi isolates in India

We generated dated phylogenies to reconstruct the evolutionary history and geographic spread of the two main lineages (2.2 and 4.3.1) circulating in Navi Mumbai. Our analysis estimated the most recent common ancestor (tMRCA) of all *S*. Typhi H58 (4.3.1) isolates in India existed around 36 years ago (1986) and was first introduced in Navi Mumbai between 1989 and 2000. Phylogeographic reconstruction indicated that the most common origin of H58 isolates observed in Navi Mumbai were Vellore (n = 8) and Mumbai (n = 7). We also identified frequent transfers between Navi Mumbai and Mumbai (n = 9). We also predicted that ciprofloxacin-resistant triple mutant isolates were introduced into Navi Mumbai on at least 12 different occasions. Two ceftriaxone-resistant H58 (4.3.1.2) isolates from our collection were genetically identical to an earlier previously *S*. Typhi isolate carrying the *bla*_SHV-12_ gene described in Eastern India (Kolkota). Our analysis also showed the ceftriaxone-resistant *S*. Typhi from Navi Mumbai were phylogenetically distant (8-45 SNPs) from previously documented ceftriaxone-resistant *S*. Typhi from Mumbai.

Phylogeographic reconstruction of the major non-H58 lineage circulating in Navi Mumbai (genotype 2.2) indicates that its ancestors were circulating in Vellore and were introduced in Navi Mumbai between 2007-2014 (Figure 2). The distribution of isolates and tree topology are consistent with at least three different transfer events of isolates from other locations to Navi Mumbai (2008-2013), followed by ongoing local expansion of a fluoroquinolone non-susceptible clade carrying a single point mutation (S83F) in QRDR region (*gyrA*).

**Figure 2.**
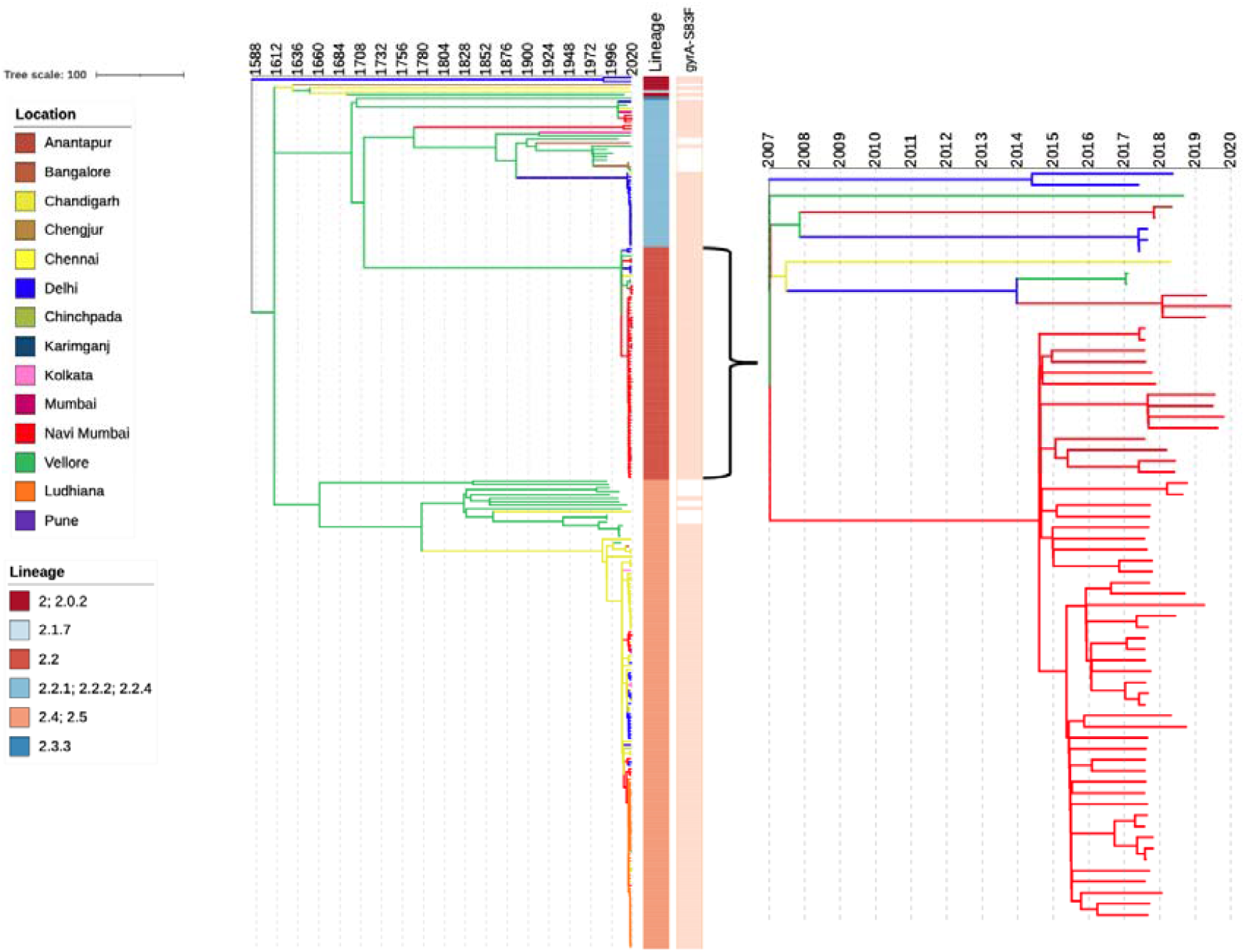
Phylogeography and expansion of *S*. Typhi lineage 2.2. Timed phylogenetic tree of genotype 4.3.1 *S*. Typhi isolates. The branch lengths are scaled in years and are colored according to the location of the most probable ancestor of descendant nodes. The scale bar indicates nucleotide substitutions per site.

**Figure 3.**
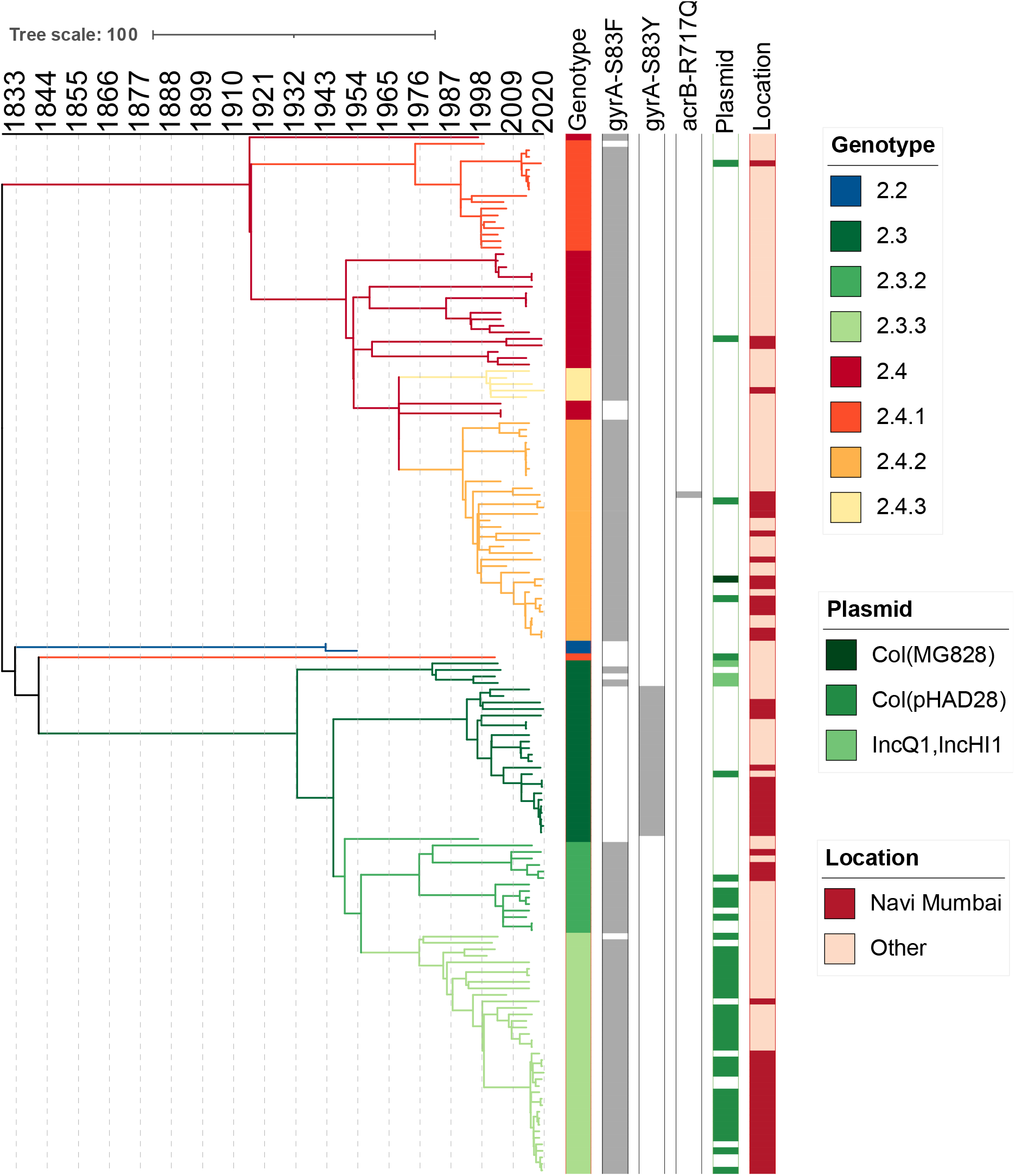
Timed phylogenetic tree of *S*. Paratyphi A isolates. The branch lengths are scaled in years and are colored according to the genotypes. The scale bar indicates nucleotide substitutions per site.

### Phylogenetic structure of S. Paratyphi A isolates in India

We identified seven different genotypes among the *S*. Paratyphi A isolates (Figure 3). All genomes from Navi Mumbai have been assigned to secondary clades 2.3 and 2.4. Genotype 2.3.3 (37.1%; 20/54), was the most common, followed by 2.3 (24.1%; 13/54), and 2.4.2 (24.1%; 13/54). To place the Navi Mumbai isolates in context, we constructed a whole genome dated phylogeny including other *S*. Paratyphi A previously sequenced in India. We estimated the tMRCA for all *S*. Paratyphi A in India existed around 1828-1832 and was first introduced to Navi Mumbai around 1992-1998. In addition, we identified at least 11 recent introductions of different genotypes to Navi Mumbai in the last 15 years.

## Discussion

Our data provide a historical insight into the composition of the circulating population structure of *S*. Typhi and *S*. Paratyphi A in India and contextualization of Navi Mumbai strains on a regional level. The close genetic relatedness of *S*. Typhi isolated in India, including the antimicrobial-resistant clades, indicates inter-regional transmission and suggests that enteric fever prevention strategies require a coordinated approach between these sites. We identified numerous transfers of H58 *S*. Typhi organisms from Mumbai to Navi Mumbai. Our observations are consistent with previous global phylogeography analysis that identified India as an important hub for the emergence and spread of antimicrobial-resistant *S*. Typhi clones (6). In contrast to the H58 analysis, phylogeography reconstruction of the major non H58 lineage (genotype 2.2) in Navi Mumbai, revealed that most typhoid cases were resulted from a recent introduction accompanied by local expansion, rather than long-term persistence.

We found that spatial distance between households of typhoid cases was strongly predictive of the probability of genetic clustering, a finding which was robust to the genetic distance threshold used to define clustering. Up to 5 kilometers, each kilometer was associated with 35% decreased odds of genetic clustering. Viewed another way, isolates from individuals living one kilometer from another had a 5.6-fold increased odds of being in a genetic cluster compared with those from individuals living 5 kilometers from one another, suggesting that much of typhoid transmission is local. The spatial scale of transmission likely varies between different communities, such that similar studies are needed elsewhere. Such findings can have important implications for informing decisions about the cluster size of randomized trials of vaccination or water, sanitation, and hygiene interventions for control of typhoid.

Our findings showed that the *S*. Paratyphi A population from Navi Mumbai is diverse, and we observe close clustering with isolates from other regions. Although *S*. Paratyphi A lineages emerged in India around 191 years ago, our analysis identified multiple recent introductions of FQNS isolates in Navi Mumbai. The frequent transfer events and high level of fluroquinolone resistance demonstrated by the different *S*. Paratyphi A genotypes is of great concern especially due the lack of an *S*. Paratyphi A vaccine which limits prevention options and highlight the importance of genomic surveillance to track the evolution of this pathogen and monitor its transmission.

The management of enteric fever is challenging due to the emergence of antibiotic-resistant *S*. Typhi strains and their changing resistance profiles (35). Cephalosporins and azithromycin are currently the first-line treatment for enteric fever in the majority of South Asian settings (12). Recent reports have established the emergence of third-generation cephalosporin resistant *S*. Typhi in India (12,36,37). In our study ceftriaxone resistance was linked to the acquisition of a IncX3 plasmid carrying the ESBL gene *bla*_SHV-12._ In general, 4.3.1.2 isolates harboring *bla*_SHV-12_ were located in independent branches of the phylogenetic tree. These data are consistent with independent acquisitions of the resistant plasmid within genotype 4.3.1.2, suggesting that the ceftriaxone resistant *S*. Typhi isolates from India have evolved independently from respective geographical locations.

The emergence and expansion of antimicrobial resistant lineages in *S*. Typhi are manly driven by antibiotic usage and selective pressure (38). This is supported by the emergence and regional dominance of H58 (4.3.1.2) QRDR triple mutant in India which was associated with high fluoroquinolone exposure (13). Previous studies reported that H58 lineage II strains with triple QRDR mutations developed cephalosporin resistance by acquiring resistance plasmids such as IncX3 (*bla*_SHV-12_) (12). The emergence and dissemination of these high-risk clones in India could lead to large outbreaks and international spread as previously observed in XDR *S*. Typhi strains carrying the ESBL gene *bla*_CTX-M-15_ in Pakistan (11).

While our findings supplement our understanding of enteric fever in an endemic setting, our study is limited by the sample of isolates available for analysis, which was small and reflects sampling of local cases. In addition, the available genomes from India might not have broad representativeness across geographic location or time. These circumstances demonstrate the importance of local laboratory and genomic surveillance in endemic regions such as Navi Mumbai to monitor the ongoing evolution of antimicrobial resistance and the impact of control strategies such as vaccination programs in India.

Our findings show that the control of enteric fever in India and South Asia requires a coordinated strategy given the inter-regional transmission of different genotypes, suggesting country-wide circulation. The emergence and transmission of high-risk lineages such as QRDR triple mutant and ceftriaxone-resistant organisms in settings with high burden of typhoid fever calls for active surveillance. The implementation of Vi conjugate vaccines appears as an essential measure to control enteric fever, but elimination will require immunization be accompanied by improvements in water sanitation and hygiene.

## Data Availability

All data produced in the present work are contained in the manuscript

## Disclaimer

The findings and conclusions in this report are those of the authors and do not necessarily represent the official position, policies, or views of the United States Centers for Disease Control and Prevention, or the World Health Organization.

## References

1. Marchello CS, Hong CY, Crump JA. Global Typhoid Fever Incidence: A Systematic Review and Meta-analysis. Clin Infect Dis [Internet]. 2019 Mar 7;68(Suppl 2):S105–16. Available from: https://pubmed.ncbi.nlm.nih.gov/30845336

2. Stanaway JD, Reiner RC, Blacker BF, Goldberg EM, Khalil IA, Troeger CE, et al. The global burden of typhoid and paratyphoid fevers: a systematic analysis for the Global Burden of Disease Study 2017. Lancet Infect Dis. 2019;19(4):369–81.

3. Antillón M, Warren JL, Crawford FW, Weinberger DM, Kürüm E, Pak GD, et al. The burden of typhoid fever in low- and middle-income countries: A meta-regression approach. PLoS Negl Trop Dis. 2017;

4. Ochiai RL, Acosta CJ, Danovaro-Holliday MC, Baiqing D, Bhattacharya SK, Agtini MD, et al. A study of typhoid fever in five Asian countries: Disease burden and implications for controls. Bull World Health Organ. 2008;

5. Park SE, Pham DT, Boinett C, Wong VK, Pak GD, Panzner U, et al. The phylogeography and incidence of multi-drug resistant typhoid fever in sub-Saharan Africa. Nat Commun. 2018;

6. Wong VK, Baker S, Pickard DJ, Parkhill J, Page AJ, Feasey NA, et al. Phylogeographical analysis of the dominant multidrug-resistant H58 clade of Salmonella Typhi identifies inter-and intracontinental transmission events. Nat Genet. 2015;

7. Karkey A, Thwaites GE, Baker S. The evolution of antimicrobial resistance in Salmonella Typhi. Current Opinion in Gastroenterology. 2018.

8. Andrews JR, Qamar FN, Charles RC, Ryan ET. Extensively Drug-Resistant Typhoid — Are Conjugate Vaccines Arriving Just in Time? N Engl J Med [Internet]. 2018 Oct 12;379(16):1493–5. Available from: https://doi.org/10.1056/NEJMp1803926

9. Britto CD, Dyson ZA, Mathias S, Bosco A, Dougan G, Jose S, et al. Persistent circulation of a fluoroquinolone-resistant Salmonella enterica Typhi clone in the Indian subcontinent. J Antimicrob Chemother. 2020;

10. Manu P. Third Generation Cephalosporins for Typhoid Fever. Am J Ther [Internet]. 2016;23(5). Available from: https://journals.lww.com/americantherapeutics/Fulltext/2016/09000/Third_Generation_Cephalosporins_for_Typhoid_Fever.1.aspx

11. Klemm EJ, Shakoor S, Page AJ, Qamar FN, Judge K, Saeed DK, et al. Emergence of an extensively drug-resistant Salmonella enterica serovar typhi clone harboring a promiscuous plasmid encoding resistance to fluoroquinolones and third-generation cephalosporins. MBio. 2018;

12. Argimón S, Nagaraj G, Shamanna V, Sravani D, Vasanth AK, Prasanna A, et al. Circulation of Third-Generation Cephalosporin Resistant Salmonella Typhi in Mumbai, India. Clin Infect Dis [Internet]. 2021 Oct 9;ciab897. Available from: https://doi.org/10.1093/cid/ciab897

13. Britto CD, Wong VK, Dougan G, Pollard AJ. A systematic review of antimicrobial resistance in Salmonella enterica serovar Typhi, the etiological agent of typhoid. PLoS Negl Trop Dis [Internet]. 2018 Oct 11;12(10):e0006779–e0006779. Available from: https://pubmed.ncbi.nlm.nih.gov/30307935

14. World Health Organization. Typhoid vaccines: WHO position paper, March 2018 – Recommendations. Vaccine [Internet]. 2019;37(2):214–6. Available from: https://www.sciencedirect.com/science/article/pii/S0264410X18304912

15. Date K, Shimpi R, Luby S, n R, Haldar P, Katkar A, et al. Decision Making and Implementation of the First Public Sector Introduction of Typhoid Conjugate Vaccine-Navi Mumbai, India, 2018. Clin Infect Dis [Internet]. 2020 Jul 29;71(Suppl 2):S172–8. Available from: https://pubmed.ncbi.nlm.nih.gov/32725235

16. India RG and CC of. 2011 Census of India [Internet]. 2011 [cited 2022 Aug 12]. Available from: https://www.census2011.co.in/census/city/368-navi-mumbai.html

17. Andrews S. Babraham Bioinformatics - FastQC A Quality Control tool for High Throughput Sequence Data. Soil. 1973.

18. Ewels P, Magnusson M, Lundin S, Käller M. MultiQC: Summarize analysis results for multiple tools and samples in a single report. Bioinformatics. 2016;

19. Wood DE, Lu J, Langmead B. Improved metagenomic analysis with Kraken 2. Genome Biol. 2019;

20. Yoshida CE, Kruczkiewicz P, Laing CR, Lingohr EJ, Gannon VPJ, Nash JHE, et al. The salmonella in silico typing resource (SISTR): An open web-accessible tool for rapidly typing and subtyping draft salmonella genome assemblies. PLoS One. 2016;

21. Inouye M, Dashnow H, Raven LA, Schultz MB, Pope BJ, Tomita T, et al. SRST2: Rapid genomic surveillance for public health and hospital microbiology labs. Genome Med. 2014;

22. Langmead B, Salzberg S. Bowtie2. Nat Methods. 2013;

23. Li H, Handsaker B, Wysoker A, Fennell T, Ruan J, Homer N, et al. The Sequence Alignment/Map format and SAMtools. Bioinformatics. 2009;

24. Wong VK, Baker S, Connor TR, Pickard D, Page AJ, Dave J, et al. An extended genotyping framework for Salmonella enterica serovar Typhi, the cause of human typhoid. Nat Commun. 2016;

25. Croucher NJ, Page AJ, Connor TR, Delaney AJ, Keane JA, Bentley SD, et al. Rapid phylogenetic analysis of large samples of recombinant bacterial whole genome sequences using Gubbins. Nucleic Acids Res. 2015;

26. Zhemin Z, Angela M, François-Xavier W, Camille B, Satheesh N, John W, et al. Transient Darwinian selection in Salmonella enterica serovar Paratyphi A during 450 years of global spread of enteric fever. Proc Natl Acad Sci [Internet]. 2014 Aug 19;111(33):12199–204. Available from: https://doi.org/10.1073/pnas.1411012111

27. Day MR, Doumith M, Do Nascimento V, Nair S, Ashton PM, Jenkins C, et al. Comparison of phenotypic and WGS-derived antimicrobial resistance profiles of Salmonella enterica serovars Typhi and Paratyphi. J Antimicrob Chemother [Internet]. 2018 Feb 1;73(2):365–72. Available from: https://doi.org/10.1093/jac/dkx379

28. Tanmoy AM, Hooda Y, Sajib MSI, Silva KE da, Iqbal J, Qamar FN, et al. Paratype: A genotyping framework and an open-source tool for &lt;em&gt;Salmonella&lt;/em&gt; Paratyphi A. medRxiv [Internet]. 2021 Jan 1;2021.11.13.21266165. Available from: http://medrxiv.org/content/early/2021/11/15/2021.11.13.21266165.abstract

29. Warren JL, Chitwood MH, Sobkowiak B, Crudu V, Colijn C, Cohen T. Spatial modeling of dyadic genetic relatedness data: Identifying factors associated with M. tuberculosis transmission in Moldova. In 2021.

30. Chattaway MA, Dallman TJ, Larkin L, Nair S, McCormick J, Mikhail A, et al. The Transformation of Reference Microbiology Methods and Surveillance for Salmonella With the Use of Whole Genome Sequencing in England and Wales. Front Public Heal. 2019;7(November):1–12.

31. Stamatakis A. RAxML version 8: A tool for phylogenetic analysis and post-analysis of large phylogenies. Bioinformatics. 2014;

32. Letunic I, Bork P. Interactive Tree Of Life (iTOL) v4: recent updates and new developments. Nucleic Acids Res. 2019;

33. Volz EM, Frost SDW. Scalable relaxed clock phylogenetic dating. Virus Evol [Internet]. 2017 Jul 1;3(2). Available from: https://doi.org/10.1093/ve/vex025

34. Thanh DP, Karkey A, Dongol S, Thi NH, Thompson CN, Rabaa MA, et al. A novel ciprofloxacin-resistant subclade of h58. Salmonella typhi is associated with fluoroquinolone treatment failure. Elife. 2016;

35. Carey ME, MacWright WR, Im J, Meiring JE, Gibani MM, Park SE, et al. The Surveillance for Enteric Fever in Asia Project (SEAP), Severe Typhoid Fever Surveillance in Africa (SETA), Surveillance of Enteric Fever in India (SEFI), and Strategic Typhoid Alliance Across Africa and Asia (STRATAA) Population-based Enteric Fever St. Clin Infect Dis [Internet]. 2020 Jul 29;71(Supplement_2):S102–10. Available from: https://doi.org/10.1093/cid/ciaa367

36. Samajpati S, Pragasam AK, Mandal S, Balaji V, Dutta S. Emergence of ceftriaxone resistant Salmonella enterica serovar Typhi in Eastern India. Infect Genet Evol [Internet]. 2021;96:105093. Available from: https://www.sciencedirect.com/science/article/pii/S1567134821003919

37. Jacob JJ, Pragasam AK, Vasudevan K, Veeraraghavan B, Kang G, John J, et al. Salmonella Typhi acquires diverse plasmids from other Enterobacteriaceae to develop cephalosporin resistance. Genomics [Internet]. 2021/05/12. 2021 Jul;113(4):2171–6. Available from: https://pubmed.ncbi.nlm.nih.gov/33965548

38. Dyson ZA, Klemm EJ, Palmer S, Dougan G. Antibiotic resistance and typhoid. Clin Infect Dis. 2019;

