## Supplementary Figure 1 for "Population structure and antimicrobial resistance patterns of *Salmonella* Typhi and Paratyphi A amid a phased municipal vaccination campaign in Navi Mumbai, India"

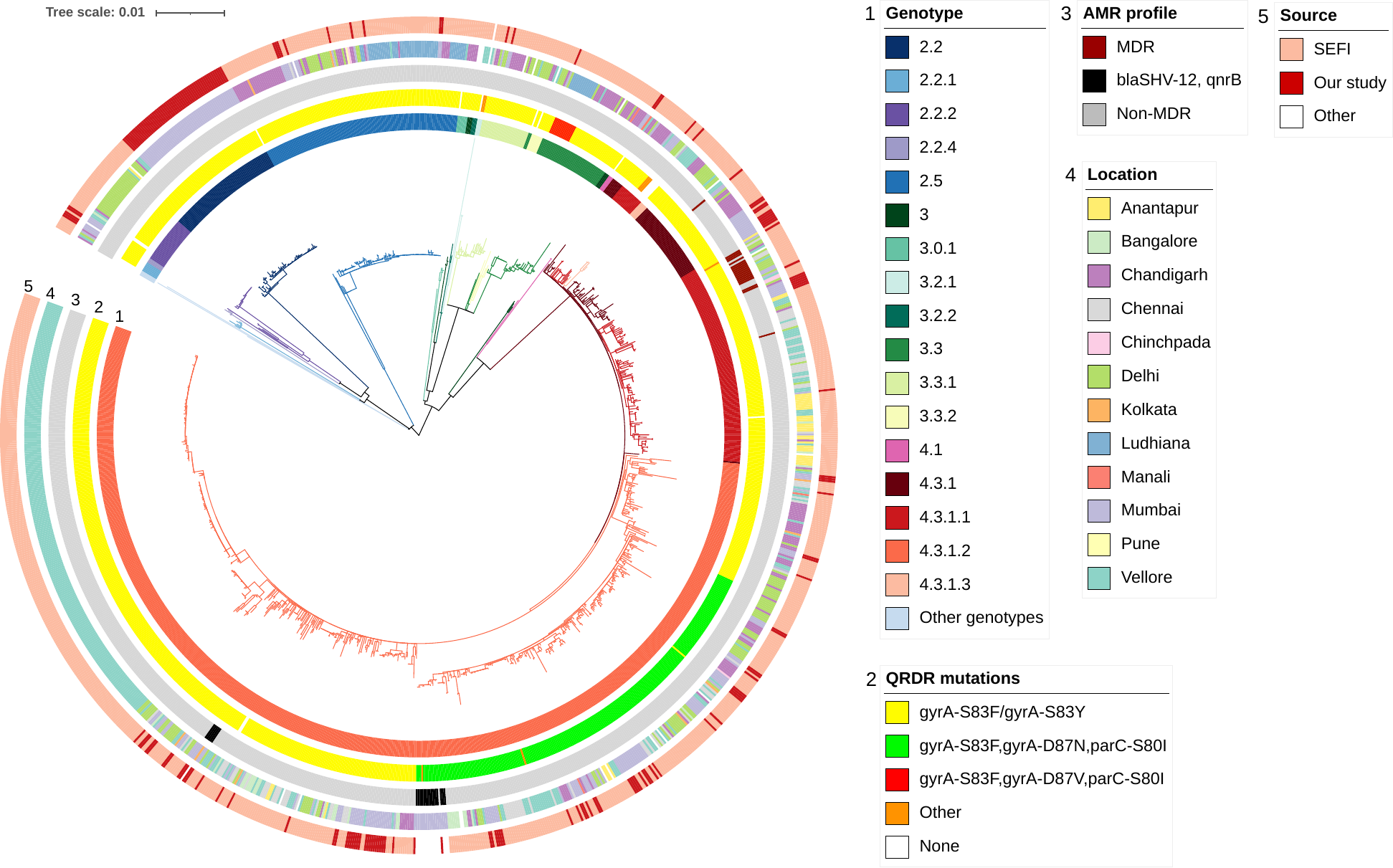

### **Figure S1.** Maximum likelihood tree of 1,533 *S*. Typhi isolates from India. Inner ring indicates the genotypes. The second ring indicates QRDR mutations profiles. Third ring indicates the antimicrobial resistance profile. Fourth ring indicates city of isolation, and the outer ring indicates the source of the study. The scale bar indicates nucleotide substitutions per site.
